# “Musculoskeletal Pathologies Affecting Shoulder Girdle: A Systematic Review with Anatomical and Radiological Validation”

**DOI:** 10.1101/2024.01.20.24301508

**Authors:** Sundip Hemant Charmode, Sudhir Kushwaha, Abhishek Kumar Mishra, Nirav Nimavat, Lalit Ratanpara, Simmi Mehra, Satish Kumar Ravi

**Author notes:** **Corresponding author:** Name: Charmode Sundip Hemant, Institution: All India Institute of Medical Sciences, Rajkot, Department : Anatomy, Address: Department of Anatomy, AIIMS Rajkot, Gujarat-360110,; 9633239685. **Dissemination history (where applicable):** This article has not been presented anywhere or the content of this article has not been published anywhere before. **Section of the journal to which the article is addressed:** Systematic Review Article. **Author contributions:** Dr. Sundip Charmode initiated the idea, designed, and drafted the article. Dr. Lt. Col. Abhishek Kumar Mishra contributed to data acquisition, analysis, and interpretation of acquired data. Dr. Sudhir Kushwaha, Dr. Lalit Ratanpara and Dr. Nirav Nimavat contributed to analysis, and interpretation of acquired data. Dr. Kumar Satish Ravi and Dr. Simmi Mehra reviewed the manuscript and gave valuable suggestions.

## Abstract

**Introduction:** Musculoskeletal pathologies affecting shoulder girdle joints, other than gleno-humeral joint such as acromioclavicular joint arthritis, tendonitis, subacromial bursitis, subdeltoid bursitis, and osteochondritis dissecans, are relatively rare. In Middle East and Asian countries, public health services are generally availed by a large number of patients in primary health centres, rural hospitals, and district hospitals, but the scarcity of specialist orthopaedic surgeons in such hospitals usually leads to misdiagnosis of rare musculoskeletal pathologies, which may result in complications and morbidity. Objectives: To determine the presentation, progression, diagnostic evaluation, and treatment of musculoskeletal pathologies affecting the shoulder girdle and develop an algorithm to screen such pathologies.

**Methods:** A systematic literature search of four medical databases (PubMed, Scopus, Web of Science and Google Scholar) was conducted, from 1^st^ January 1950 to 31^st^ December 2022. Studies (case reports, prospective studies, review articles) reporting pathological conditions affecting shoulder girdles, with a focus on clinical presentation, physical examination tests required for diagnostic evaluation, and management, were included. The relevant data was extracted from the selected studies and tabulated for analysis.

**Results and Discussion:** Seventeen studies were included in the final analysis. Several case reports, case series and review articles showed that very few musculoskeletal conditions can be correctly diagnosed based on only physical examination tests. Radiological and anatomical basis of each of the cases were discussed.

**Conclusion:** An algorithm was prepared to help diagnose shoulder girdle pathologies based on clinical presentation and examination findings.

**Key message:** An algorithm was developed which will facilitate precise diagnosis of musculoskeletal conditions affecting shoulder girdle. The musculoskeletal pathologies of shoulder girdle were categorised based on the modalities that led to their confirmatory diagnosis, and the modes management that resulted in complete functional recovery.

## INTRODUCTION

Musculoskeletal pathologies affecting the pectoral girdle, constitute commonly the degenerative joint diseases such as glenohumeral joint osteoarthritis (GHOA), which involves the articular cartilage, subchondral, periarticular bone, articulating bones, and periarticular soft tissues of the glenohumeral joint (GHJ) as well as surrounding structures. The prevalence of GHOA is 16%–20% in the middle-aged and elderly populations^1^. In the younger population, the prevalence is influenced by inheritance, sex, obesity, joint-cavity infection, history of trauma, or occupational history of performing heavy labor^2^. GHOA is not the most frequent cause of shoulder discomfort and is frequently diagnosed after excluding more prevalent shoulder disorders^3^. This systematic review aimed to describe and discuss the presentation, progression, diagnostic evaluation, and treatment of musculoskeletal pathologies affecting the shoulder girdle and develop an algorithm to screen such pathologies.

## METHODS

### Prospero protocol

To reduce the risk of bias in the study, the authors prepared and prospectively registered a research review protocol in PROSPERO at the Center for Reviews and Dissemination, University of York (CRD42023475189). The review protocol can be accessed at https://www.crd.york.ac.uk/prospero/.

### Search strategy

According to the Preferred Reporting Items for Systematic Reviews and Meta-Analyses (PRISMA), an initial online search (scoping review) of articles published in four medical databases (PubMed, Scopus, Web of Science, and Google Scholar) from 1950 to 2022 was conducted. The search was performed using the following MeSH terms: “osteoarthritis,” “adhesive capsulitis,” “glenohumeral joint,” acromioclavicular joint,” “bursitis,” “rotator cuff tear,” “tendinopathy,” “fractures of upper end of humerus,” “osteochondritis dissecans,” “rheumatoid arthritis,” “bicipital tendonitis,” and “glenoidal labrum tears.”

### Eligibility criteria

The inclusion criteria were complete articles (free full text) published between January 1, 1950 and December 31, 2022; articles involving human subjects; articles published by both foreign and Indian authors; and articles focusing on arthropathy of the GHJ, acromioclavicular joint (ACJ) arthritis, rotator cuff tear, glenoidal labrum injury, subacromial bursitis, tendinopathy/tendinosis, subdeltoid bursitis, rheumatoid arthritis (RA), osteochondritis dissecans (OCD) of the greater and lesser humeral tubercles, and adhesive capsulitis. Furthermore, the inclusion criteria included isolated muscle tendinosis of the pectoral girdle region and fractures of the upper end of the humerus.

The exclusion criteria were articles published before January 1, 1950 and after December 31, 2022; articles accessible through abstracts only; and publications focusing on topics other than the abovementioned conditions. Moreover, non-English articles were excluded. Non-peer-reviewed publications, such as Letters, Commentaries, Short communication, and newsletters, were also excluded. Articles (original, review, case series, case reports, published, and preprints) that satisfied the inclusion and exclusion criteria were included in this review. The Institutional Ethics Committee (IEC) was informed regarding the aim of this study, and the need for IEC approval was waived as that this review only contained patient data that are freely available in the public domain.

### Study selection

After applying the inclusion and exclusion criteria, two authors (SC and AM) independently assessed the titles and abstracts of all articles found during the initial search, after which eligible articles were shortlisted. These articles were downloaded and independently examined based on the data extraction checklist prepared by both authors for relevant data. The authors of the articles and methods of the investigations were not concealed from the reviewers. Any differences were resolved through discussion or involvement of a third reviewer (SK). The data relevant for our study and the excluded data are described in the checklist shown in Table 1. We selected and incorporated the articles that contained this information.

### Data extraction process

The authors formulated a “data extraction form” to extract relevant data. This form was pilot-tested using ten sample articles before actual use. Two reviewers (SC and LR) worked separately to collect data from the selected articles. Disagreements were resolved through discussion and involvement of a third independent reviewer (SK). The following features of the study were collected: (i) name of the pathological condition, (ii) presenting signs and symptoms, (iii) patient’s age, (iv) medical or surgical illness history, (v) anatomical basis of the condition, and (vi) histological basis of the condition. The following information about the pathology of the shoulder girdle was collected: (i) initial diagnosis at first visit, (ii) investigations conducted to confirm the diagnosis, (iii) local examination performed to identify the involved musculoskeletal structures and their findings, recommended treatment, recommended mode of physiotherapy, follow-up, return to previous functional level, and history of any recurrence.

### Risk of bias

While selecting the articles, the following eligibility criteria were used to calibrate the assessments between the two reviewers at the end of every five articles. If a high level of agreement (>90%) was not achieved between the reviewers, they discussed their points of disagreement and reviewed (and if required, revised) the inclusion criteria. Two authors used the data extraction form to review five selected articles separately. After a review of every five articles, they discussed any discrepancies in the collected data and determined if there was any need to modify the data extraction form. In this manner, the reviewers assessed the risk of bias. Subsequently, the risk of bias in systematic reviews (ROBIS) tool was used to assess the risk of bias in our review^4^.

## RESULTS

### Literature search

The initial online search using “arthropathy of shoulder joint” as the MeSH term resulted in 19,653 articles, and after using “arthropathy of shoulder girdle” as the MeSH term, the search provided 450 articles. After applying various search filters, such as human involvement, English language, publication dates between 1950 and 2022, and free full text, the number of publications was reduced to 152. Duplicate articles (n = 23) were deleted. Two reviewers (SC and LR) independently assessed each article. Seventy-seven articles were excluded due to irrelevant text outside of the study scope. After applying the eligibility criteria, 52 articles were shortlisted. After a citation search, 17 articles were additionally included, which were separately reviewed again by the same two authors. After a review of 69 shortlisted articles by the designated authors (SC and LR) using the data extraction checklist and subsequent discussion, 17 articles were finally included in this study. **“**Figure 1 Preferred Reporting Items for Systematic Reviews and Meta-Analyses (PRISMA) flowchart with its step-by-step literature search and consideration/rejection procedure”.

**Figure 1.**
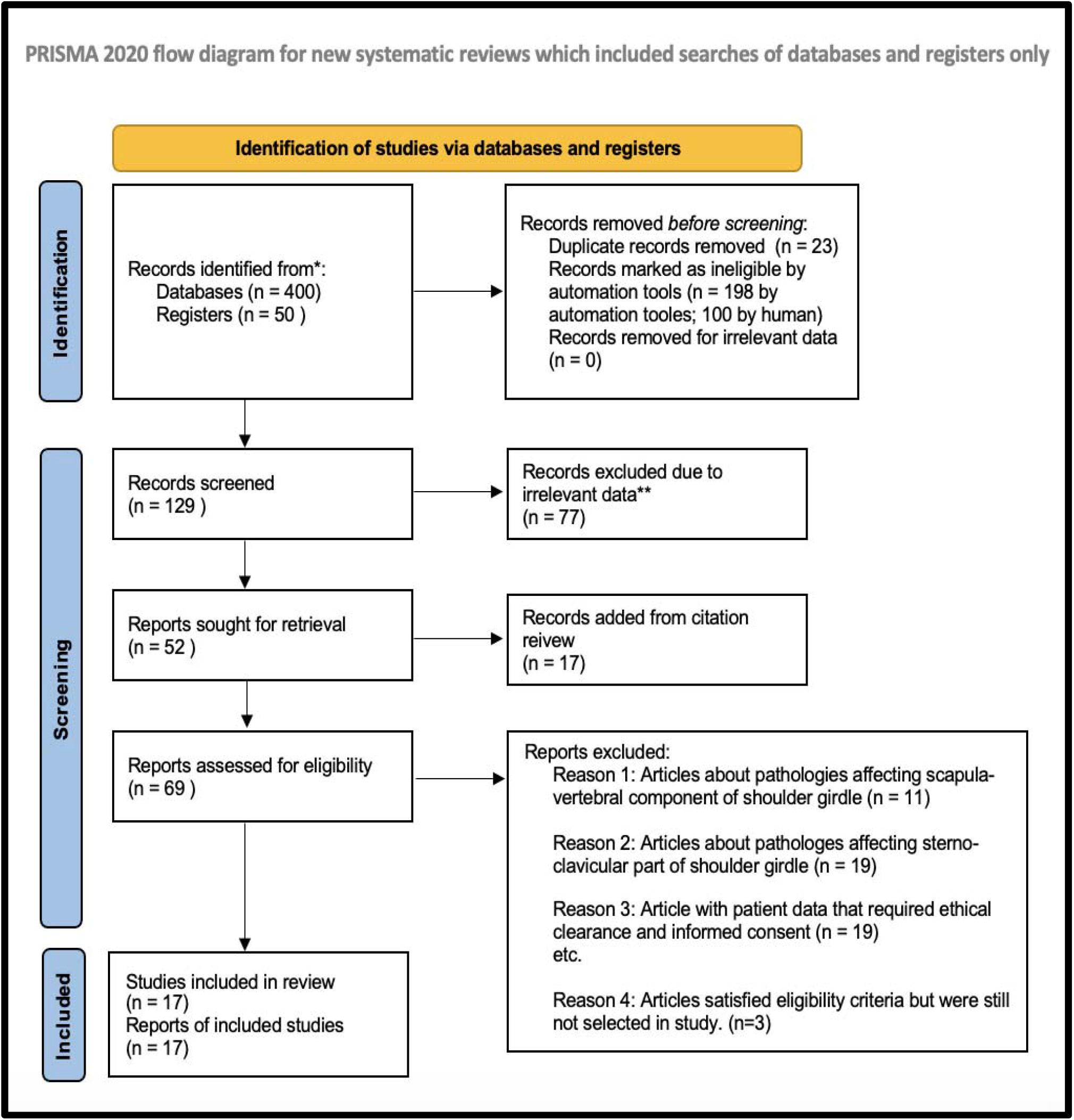
shows the Preferred Reporting Items for Systematic Reviews and Meta-Analyses (PRISMA) flowchart, which shows the step-by-step literature search and consideration/rejection procedure.

### Studies that met the inclusion criteria but were excluded from the study

Three articles were shortlisted during the online search, based on the inclusion criteria but were not selected in the study. The studies were Logli AL (2021), Cole BJ (2007), and Labriola JE (2005). Logli AL and his colleagues proposed the impact of angulation defects of humeral head and glenoid bone on progression of gleno-humeral joint arthritis and classified the gleno-humeral joint arthritis into eccentric and concentric types [5]. This is a new concept which needs validation, so study was excluded from our review. Cole BJ and his team reviewed non-arthroplasty treatment methods of gleno-humeral joint arthritis but focused on the challenges associated with the diagnosis and management of glenohumeral arthritis in patients having other shoulder co-morbidities^6^. Labriola JE and his colleagues in 2005 studied the factors of stability and instability of shoulder girdle with respect to shoulder muscles^7^.

### Characteristic features of each selected study

Out of the seventeen selected articles, six articles were review studies, two articles were case report and review study, eight articles were case reports, and one article was of prospective study design. An algorithm based on physical examination findings was developed to screen various pathologies of the shoulder girdle (Figure 2).

**Figure 2.**
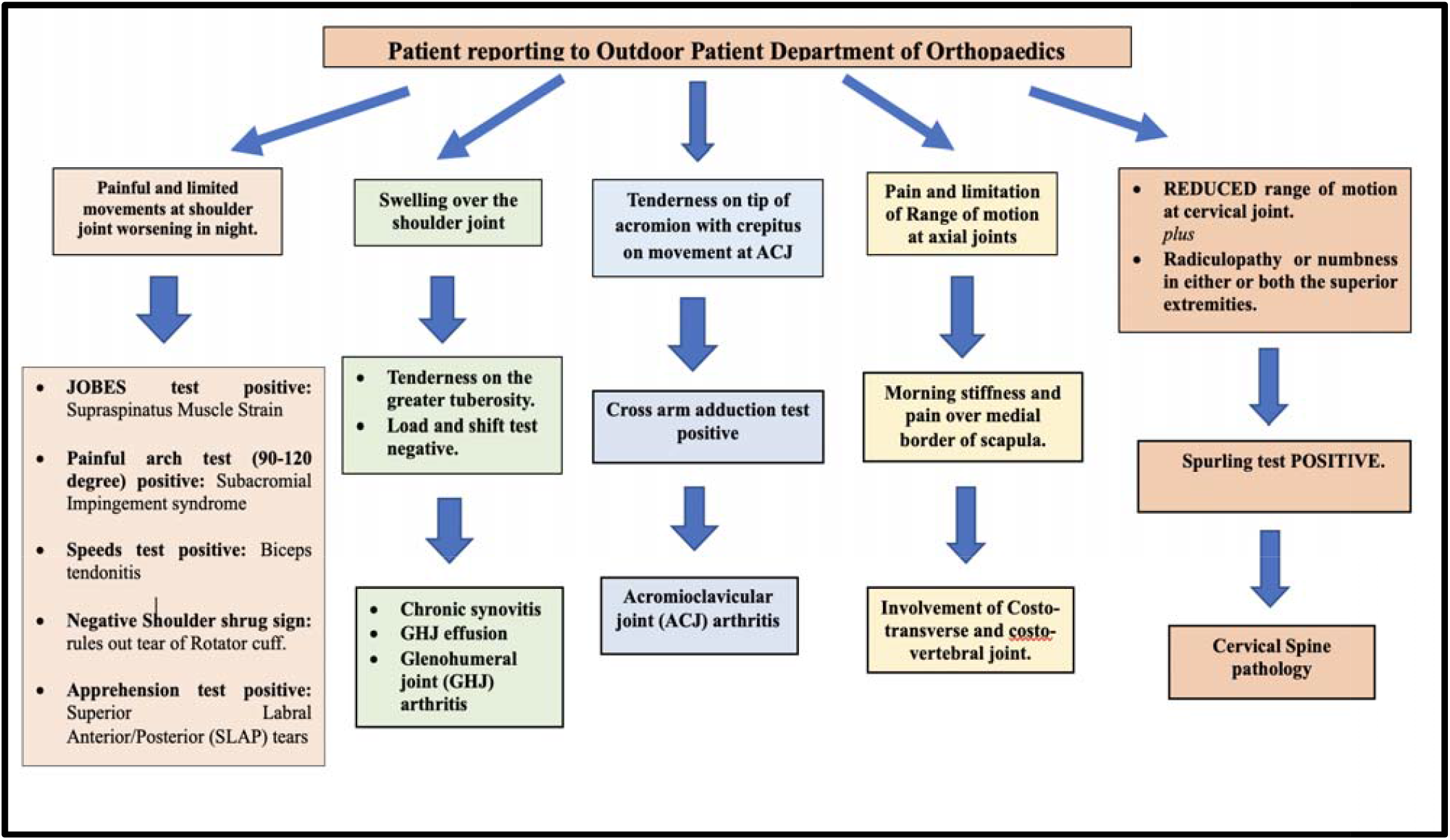
presents an algorithm formulated for screening of various pathologies of shoulder girdle based on physical examination findings. GHJ: Gleno-humeral joint; ACJ: Acromio-clavicular joint; SLAP: Superior Labral Anterior/Posterior tears; Anti-CCP: Anticyclic Citrullinated bodies.

## DISCUSSION

### Surgical anatomy of the shoulder girdle

The shoulder girdle comprises three articulating bones (distal end of the clavicle, acromion process of the scapula, and head of the humerus connected with a few ligaments and muscles) and two joints: the GHJ and ACJ. Figures 3 and 4 depict the interior and exterior of the shoulder joint, respectively^8,9^.

**Fig. 3.**
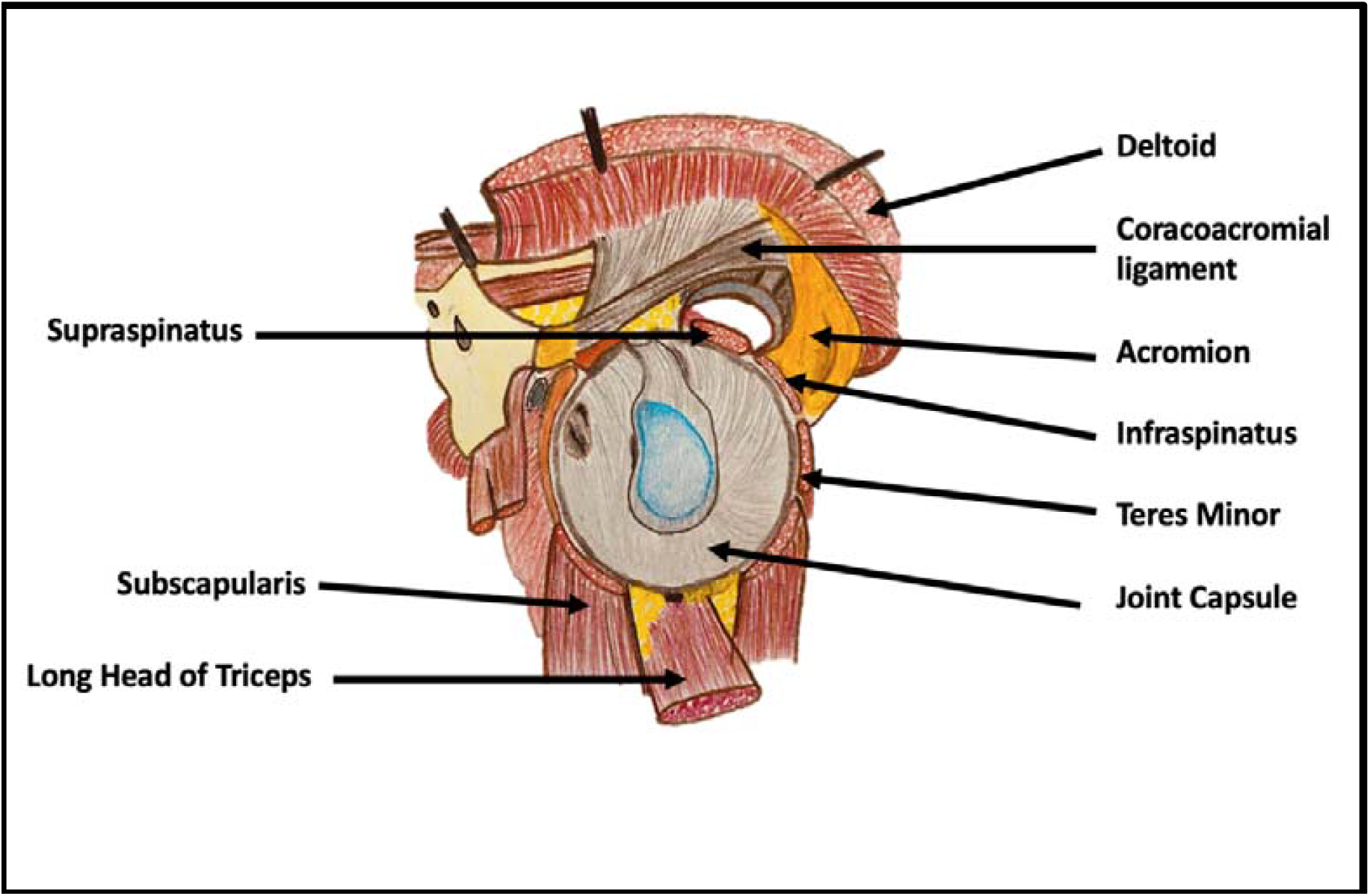
The Interior of shoulder joint, Antero-lateral aspect [8]. (Courtesy: GRAY’S Anatomy. The Anatomical Basis of Clinical Practice. London. Elsevier. 41^st^ edition. 2016. Section 6: Chapter 48. Joints. Figure 48.15. Page 809. This image was created and edited by Dr. Rishita Vala, Junior Resident, AIIMS Rajkot)

**Fig. 4.**
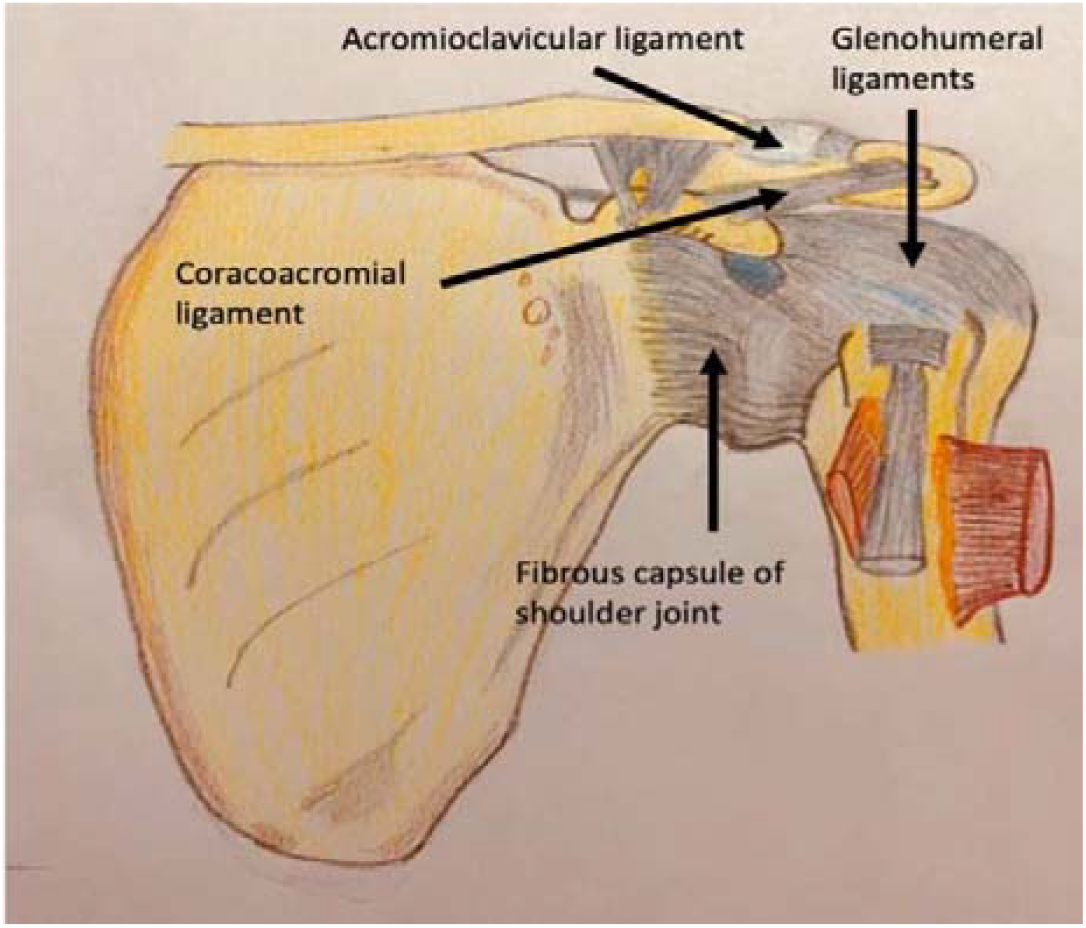
Ligaments associated with shoulder joint, anterior aspect [9]. (Courtesy: GRAY’S Anatomy. The Anatomical Basis of Clinical Practice. London. Elsevier. 41^st^ edition. 2016. Section 6: Chapter 48. Joints. Figure 48.16. Page 810. This image was created and edited by Dr. Rishita Vala, Junior Resident, AIIMS Rajkot)

### Comparative review

The findings of the current case correlated with those of the reference cases and are presented in Table 2.

#### Shoulder girdle pathologies diagnosed based on physical examination tests

Most shoulder girdle pathologies present with specific signs and symptoms of pain, tenderness, and reduced ROM, which can be identified using physical examination tests. Nejati et al. reported that greater tenderness over the coracoid process is a critical sign in conditions such as adhesive shoulder capsulitis^10^. In their scoping review, Chaudhury et al. revealed that in patients with ACJ arthritis, physical examination is enough for diagnosis as the pain is usually localized to the ACJ with no radiation^14^. Gorkiewicz reported that subacromial bursitis was diagnosed in a 38-year-old woman with a sedentary lifestyle based on an examination; a painful ROM between 60° and 120° was observed, and the speed test was positive^15^. In a case report by Goyal et al., based on a positive speed test and Neer’s test, with decreased ROM of the shoulder, subdeltoid bursitis was diagnosed in a 70-year-old woman reporting painful shoulder movements localized over the lateral aspect of a shoulder^16^.

#### Shoulder girdle pathologies that require radiological investigations for diagnosis

Imai et al. reported that a few conditions such as a rotator cuff partial or complete tear can be identified via a positive empty can test, but MRI is required to confirm the diagnosis^11^. Lewis et al. revealed that physical examination can aid in the diagnosis of this illness, but ultrasonography seems to be the most reliable approach to obtain a firm diagnosis of bicep tendinopathy^12^. Khan et al. reported that subacromial impingement syndrome was diagnosed based on a positive Neer test, but MRI was essential to confirm the diagnosis based on a degenerated and swollen supraspinatus tendon (critical zone)^13^. Maalouly et al. reported a 29-year-old man who presented with a repeated history of dislocation of a shoulder joint, wherein a glenoidal labral tear was identified based on positive apprehension test, relocation test, and Hawkins–Kennedy test. Confirmatory diagnosis was obtained via MRI and noncontrast CT of the shoulder joint, and the patient was managed via arthroscopic Bankart repair^17^. Barakat et al. reported that a fractured neck of the humerus was diagnosed in a 44-year-old man based on physical examination tests, but it required confirmation via radiological examination^18^. Jafari et al. reported a 37-year-old woman presenting with right shoulder pain after a fall 15 days ago. They revealed that even though the diagnosis of OCD was based on a positive Hawkins–Kennedy test, internal rotation against resistance, and belly press test, the diagnosis had to be further confirmed via MRI^19^. Epstein et al. conducted a review and emphasized the utility of MRI in diagnosing rupture of the coracobrachialis and short head of the biceps brachii and recommended prompt surgical reconstruction to achieve a successful outcome with no functional residual deficit^21^. In a case report, Alben et al. described that despite a positive physical examination test (painful external rotation against resistance) in a 52-year-old woman presenting with pain over the posterior aspect of a shoulder, MRI was eventually required to diagnose teres minor and infraspinatus tears^22^. In a case report, Spargoli described a supraspinatus tear that was identified based on physical examination, including pain on initiation of shoulder abduction and positive results of Hawkins–Kennedy and empty can test, but confirmation via MRI was needed^23^. Ishida et al. conducted a prospective study and emphasized the difficulty of diagnosing RA in patients with rotator cuff tears. The authors stated that RA should be suspected when MRI findings show rotator cuff tear with significant synovial proliferation. They highlighted the usefulness of pathological biopsy for early diagnosis of RA onset^24^.

#### Shoulder girdle pathologies that require only physical therapy for full restoration of movement

Patients with conditions such as adhesive shoulder capsulitis respond well to physical therapy (shoulder girdle exercises) for 6 months, which helps regain full ROM^10^. Subacromial impingement syndrome is treated with conservative management (NSAIDS, rest, ice pack, and prolonged physical therapy), with patients typically regaining full ROM within 6 months^13^.

#### Shoulder girdle pathologies that require surgical intervention

Imai et al. reported that arthroscopic repair of the rotator cuff followed by passively assisted and pendulum exercises, which is further followed by rotator cuff strengthening activities at postoperative 7 weeks, was required for full restoration^11^. Patients with ACJ arthritis are currently managed via physiotherapy initially, followed by ACJ steroid injection, which is further followed by surgery^14^. Gorkiewicz reported that when patients with subacromial bursitis were treated conservatively initially, it did not lead to improvement; therefore, a subacromial steroid injection was administered^15^. Goyal et al. demonstrated that subdeltoid bursitis was initially treated with NSAIDs and physiotherapy but later with arthroscopic excision of the bursa and lateral end of the acromion debridement^16^. Hamada et al. studied a rare case of OCD and stated that repetitive microtrauma is the cause of OCD and that the most effective treatment of OCD with a partial detached fragment is its removal and drilling^20^ Sambandam et al., explained how a patient with restricted shoulder movements required further evaluation with MRI which revealed a septic arthritis of left gleno-humeral joint for which emergency arthroscopic debridement was performed^25^. Lossos IS. et. al., in their study of eleven cases stated that earlier x rays were inconclusive, and ultrasonography was required to detect the fluid in the joint cavity^26^.

## Data Availability

All data produced in the present study are available upon reasonable request to the authors

## PROSPERO registry number

CRD42023475189

## Author contributions

SC initiated the idea and designed and drafted the article. LR contributed to data acquisition, analysis, and interpretation of the acquired data., AKM, SK and SM contributed to the analysis and interpretation of the acquired data. SKR and NN reviewed the manuscript and provided valuable suggestions.

## Acknowledgements

We extend our sincere gratitude toward our respected Executive Director, Prof. Dr. Colonel. CDS Katoch, who has constantly encouraged us to conduct quality research work.

## Summary of work done by the contributors

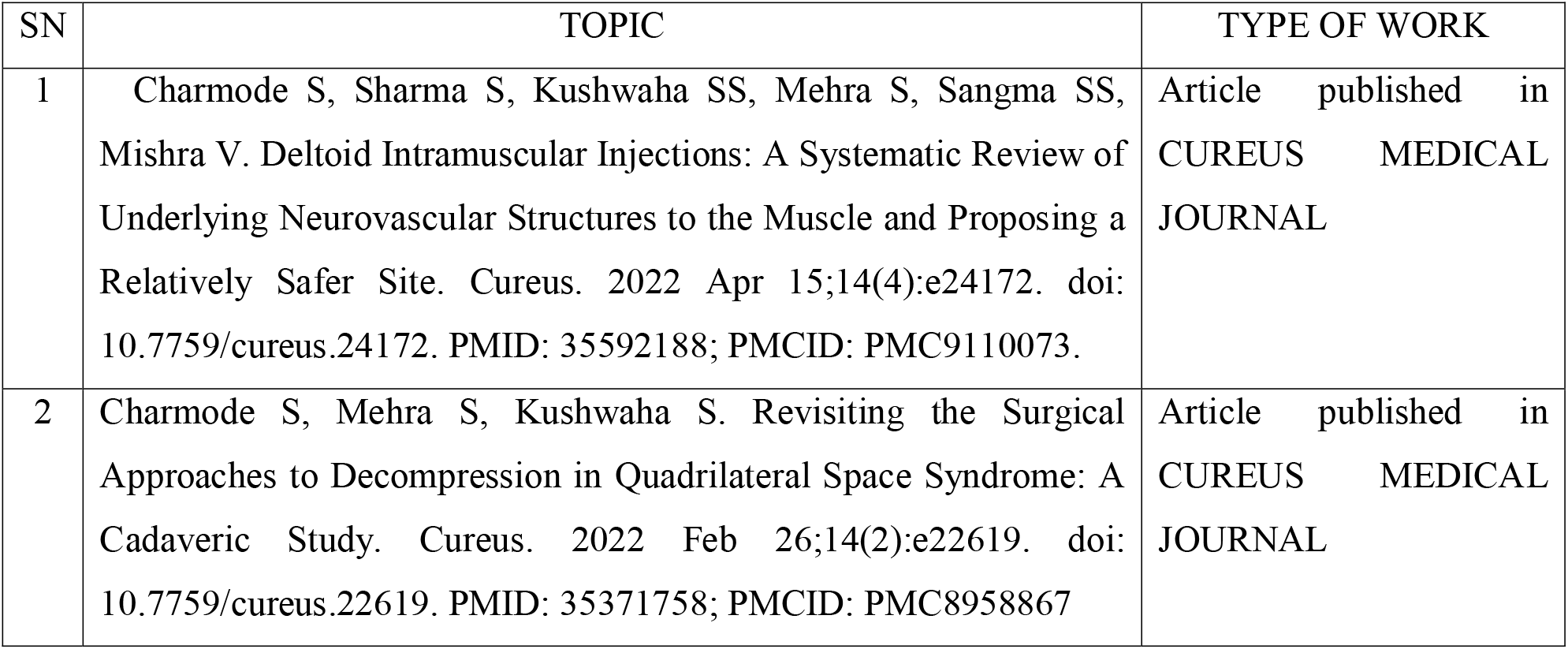
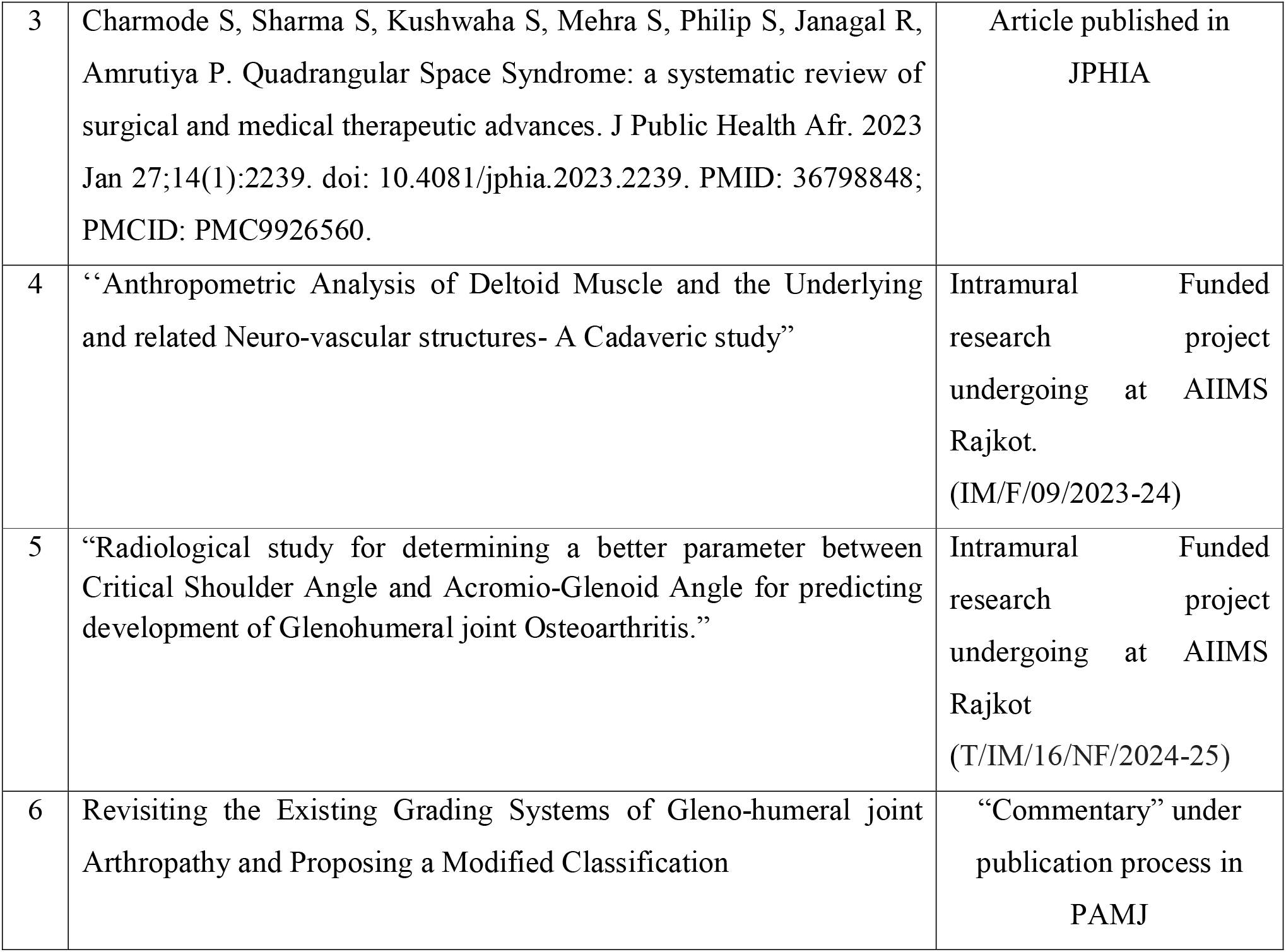

