## Supplementary material for "“Musculoskeletal Pathologies Affecting Shoulder Girdle: A Systematic Review with Anatomical and Radiological Validation”": Tables

**List of Tables**

**Table 1. Checklist for data extraction from the selected articles.**

| **Sl.**  **no** | **Data collected** | **Data ignored** |
| --- | --- | --- |
| 1 | Age group and gender of the patient. | Demographics of the patient. |
| 2 | Past medical and surgical history of the patient. | Data about the hospital, or institute where the patient was treated. |
| 3 | Presenting signs and symptoms of the patient and progression of the condition. | Data about pathologies affecting scapular -vertebral component of pectoral girdle. |
| 4 | Clinical examination of affected component of shoulder girdle with findings. | Data about pathologies affecting sterno-clavicular part of shoulder girdle. |
| 5 | Laboratory and radiological investigations advised for confirmation of diagnosis. | Data regarding any patient that required ethical clearance and informed consent was not used. |
| 6 | Title of the pathological condition diagnosed. | Review of older, current, and newer therapies of treating arthropathies of gleno-humeral joint. |
| 7 | Mode of treatment offered from the provisional till the final diagnosis. |  |
| 8 | Mode of physiotherapy offered (if applicable) |  |
| 9 | Follow up and time taken for recovery to previous functional level. |  |
| 10 | History of any recurrence. |  |

**Table 2. Comparative review with pathologies of shoulder girdle with its anatomical basis:**

| **Case no** | **Author details, year of publication and country of origin** | **Study design** | **Clinical sign/symptoms** | **Causative**  **pathologies** | **Anatomical basis of the pathology** |
| --- | --- | --- | --- | --- | --- |
| 1 | Nejati and Akbari, 2014 [9] | Case report | Painful and limited ROM at GHJ. | Adhesive capsulitis | Inflammation causes loss of synovial layer of capsule; reduced capsular volume; contracture of GH capsule; adhesion of axillary fold to itself and anatomical neck of humerus; thickened and fibrotic rotator interval; contracted coraco-humeral ligament (CHL).  Fibroblasts with ability to transform into smooth muscle phenotype (myofibroblasts) leads to contracture. |
| 2 | Imai et. al., 2021 [10] | Case report | Moderate to severe painful with limited ROM at GHJ after history of trauma. The pain is at night and at rest particularly. | Rotator cuff tear/injury | Four muscles namely supraspinatus, infraspinatus, subscapularis, and teres minor form the rotator cuff so each muscle needs to be tested. Jobe test, External Rotator lag sign, Hornblower sign and Belly press test. Supraspinatus tear is common. Partial or complete tears. Wear and tear (degenerative) or injury are the causes of tear. |
| 3 | Lewis et al., 2016 [11] | Review article | Mild anterior shoulder pain with normal ROM and no trauma history. Pain radiates to anterior aspect of arm. | Bicipital tendonitis/ tendinopathy | Inflammation of tendon around the long head of biceps in the intertubercular groove is primary biceps tendinitis (5%). 95% are accompanied with rotator cuff tear or labral tears. Pair is distinctively in the bicipital groove. |
| 4 | Khan et al., 2013 [12] | Review article | Moderate to severe pain at antero-lateral deltoid region with tenderness over acromion process. Full ROM but pain at 90^0^ flexion, 95^0^ of abduction, and worst with horizontal adduction. | Sub-acromial impingement syndrome | Inflammation and irritation of the rotator cuff tendons as they pass through the subacromial space, resulting in pain, weakness, and reduced range of motion.  Injuries vary from mild tendon inflammation (tendonitis), bursitis (inflamed bursa), calcific tendonitis (a calcium deposit forming within the tendon), tendinopathy or partial and full tears of the rotator cuff muscles. |
| 5 | Chaudhary S. et. al., 2018 (UK) | Review article | Pain over superior aspect of shoulder joint with tenderness over ACJ. May have history of trauma or may be asymptomatic. | Acromio-clavicular joint (ACJ) arthritis | Any type of injury to shoulder or overuse of ACJ or GHJ or older age leads to degeneration of articular disc/cartilage of ACJ; reduction of joint space causing painful movements. |
| 6 | Robert Gorkiewicz, 1984 (USA) | Case report | Anterior or anterolateral shoulder pain aggravates on overhead abduction. Worse at night and on sleeping on affected side. Wearing of cloth and combing hair become difficult. | Sub-acromial bursitis | Inflammation of the subacromial bursa, which causes increased fluid and collagen formation by the synovial cells of the bursa. The fluid is often rich in fibrin and can become haemorrhagic. |
| 7 | Goyal T, Nag HL, Tripathy SK; 2012 (India) | Case report | Painful shoulder movements. It may be localized to shoulder. It may be more generalized involving whole arm. | Sub-deltoid bursitis | Inflammatory change of the bursa is consistent with an increased amount of fluid and collagen formation e.g., because of excessive friction. |
| 8 | Maalouly J, Aouad D, Tawk A, Dib N, El Rassi G.; 2020 (Lebanon) | Case report | Glenoid labral tear can present in varied ways depending upon site and extent of it.  Antero-inferior tear (Bankart Lesion) presents as recurrent dislocation shoulder.  Superior Labral tears anterior to posterior (SLAP) presents as instability symptoms in lesser severity and chronic pain. Posterior tear (Reverse Bankart) may also present with instability features. | Glenoidal labral tear | Glenoidal labral is fibrous cartilage, but newer studies reveal that it is composed of dense fibrous collagen tissue. Superior and inferior labrum exhibit significant different anatomy. Superior labrum is rather loose, mobile and has a “meniscal‐like” aspect, while the inferior labrum appears rounded and more tightly attached to the glenoid rim. |
| 9 | Barakat A, Hatrick NC; 2019 (UK) | Case report | Usually, fractures affecting old and osteoporosis population with trivial trauma history.  Patient presents with pain, swelling and difficult shoulder and upper limb movements. | Fracture affecting surgical neck of humerus | Axillary nerve and branch of posterior circumflex humeral artery lies in relation to the surgical neck of humerus. |
| 10 | Jafari D, et. al., 2017 (Iran) | Case report and Review article | Pain in the shoulder joint with history of incidental injury, which brings pain to the notice of the patient. Pain aggravates during overhead movements and internal rotations. | OCD of Greater tubercle of humerus | The sub-chondral bone with or without its articular cartilage suffers lack of blood supply and undergoes degeneration. |
| 11 | S Hamada et. al., 1992 (Japan) | Review article | Pain in anterior aspect of shoulder which aggravates on forcefully flexion and forward elevation. | OCD of Lesser tubercle of humerus | The sub-chondral bone with or without its articular cartilage suffers lack of blood supply and undergoes degeneration. |
| 12 | Epstein SH, et. al., 2019 (USA) | Case report and Review article | Pain medial to glenohumeral joint line with tendency to radiate down below on flexion and elevation. Pain may aggravate on flexion. | OCD of tip of coracoid process/tendinopathy of coracobrachialis/ short head of biceps | The sub-chondral bone with or without its articular cartilage suffers lack of blood supply and undergoes degeneration. |
| 13 | Alben MG, Gambhir N, Virk MS; 2022 (USA) | Case report | Usually following exertions injury or may be with aging. Difficulty and pain in external rotation. | Isolated tendinosis of Teres minor and Infraspinatus | Inflammation or irritation of tendon of teres minor and infraspinatus due to repetitive overuse. |
| 14 | Spargoli G., 2018 (Italy) | Review article | Pain in anterolateral aspect of shoulder or proximal 1/3^rd^ of arm. Painful and difficult overhead abduction. | Isolated supraspinatus tendinosis | Inflammation or irritation of tendon of supraspinatus due to repetitive overuse. |
| 15 | Ishida K. et. al., 2021 (Japan) | Prospective study | Pain and reduced ROM shoulder joint. Difficulty and pain in all ROM of the joint. In advanced cases there may be massive inflammatory fluid accumulation in the joint. In late stages there may be deformity and destroyed joint. | Rheumatoid arthritis affecting ACJ or GHJ | Non-specific inflammation within the joint progressing to proliferative lesion within the synovium leading to joint destruction. It initiates with exudative phase involving the microcirculation and lining cells of synovium, which allows the influx of plasma proteins in the sub-synovium. This phase is followed by chronic inflammatory phase. |
| 16a and 16b | Sambandam SN, and Atturu M; 2016 (India) | Case report | Painful restricted movements at GHJ. Formation of osteophytes and enchondral ossification in the transition area of the hyaline cartilage and synovial membrane. | Glenoid-humeral osteoarthritis (GHOA) | Primary GHOA:  Articular cartilage changes: Irreversible progressive loss with hypertrophic reaction of the subchondral bone.  Humeral head: thinning/absence of cartilage, flattening, osteophyte and subchondral cyst formation.  Posterior wear of glenoid cavity  Rotator cuff tear in 5-10 % cases  Secondary GHOA: articular surface incongruities. |
|  | Lossos IS. Et. al., 1998 (Israel) | Review article |  |  |  |

ROM: Range of motion; ACJ: Acromio-clavicular joint; GHOA: Glenoid-humeral osteoarthritis; CHL: Coracohumeral ligament; OCD: Osteochondritis dissecans; GHJ: Gleno-humeral joint.
